# Quality of life and associated factors among HIV positive patients after completion of treatment for Cryptococcal meningitis

**DOI:** 10.1101/2020.06.19.20135368

**Authors:** Jonathan Kitonsa, Julius Kiwanuka, Zacchaeus Anywaine, Sheila Kansiime, Kenneth Katumba, Namirembe Aeron, Justin Beardsley, Freddie Kibengo, Alastair Gray, Pontiano Kaleebu, Jeremy Day

**Author notes:** **Corresponding author**, (JK).

## Abstract

**Background:** Cryptococcal meningitis (CCM) remains one of the leading causes of mortality among HIV infected patients. Due to factors such as the severity of CCM pathology, the quality of life (QOL) of patients post-treatment is likely to be poor. Few studies have reported on QOL of CCM patients post treatment completion. We used data collected among patients in the CryptoDex trial (ISRCTN59144167) to determine QOL and associated factors at week 10 and six months from treatment initiation.

**Methodology:** CryptoDex was a double-blind placebo-controlled trial of adjunctive dexamethasone in HIV infected adults with CCM, conducted between 2013 and 2015 in six countries in Asia and Africa. QOL was determined using the descriptive and Visual Analog Scales (VAS) of the EuroQol Five-Dimension-Three-Level (EQ-5D-3L) tool. We derived index scores, and described these and the VAS scores at 10 weeks and 6 months; and used linear regression to determine the relationship between various characteristics and VAS scores at both time points.

**Results:** Of 451 patients enrolled in the trial, 238 had QOL evaluations at week 10. At baseline, their mean age (SD) was 35.2(8.5) years. The overall mean VAS score (SD) at 10 weeks was 57.2 (29.7), increasing significantly to 72(27.4) at month six (p<0.001). The overall mean VAS score (SD) at week 10 was 57.2(29.7), increasing significantly to 72(27.4) at month six, (p<0.001). At week 10, higher VAS score was associated with absence of confusion (p=0.039), greater weight (p=0.002), and being African (p<0.001). At month six, higher VAS score remained associated with African origin (p=0.019). Higher number of inpatient days was associated with worse VAS scores at 10 weeks and 6 months (p<0.001 and p=0.006 respectively).

**Conclusion:** QOL was good among patients that had completed therapy for CCM, but below perfect. Strategies to improve QOL among CCM survivors are required.

**Lay summary:** In spite of the remarkable reduction in the incidence of Cryptococcal meningitis (CCM), with increased use of antiretroviral therapy, incidence remains unacceptably high especially in sub-Saharan Africa and Asia where more than 90% of the cases and deaths occur.

Due to factors such as the severity of CCM pathology, the quality of life (QOL) of patients post-treatment is also likely to be poor. Few studies have reported on QOL of CCM patients post treatment completion. We used data collected among patients in the CryptoDex trial (ISRCTN59144167) to determine self-perceived QOL and associated factors among 238 survivors at week 10 and 203 survivors at six months from treatment initiation.

We determined QOL using the descriptive and Visual Analog Scales (VAS) of the EuroQol Five-Dimension-Three-Level (EQ-5D-3L) tool.

We found that while self-perceived QOL was only relatively good among this cohort of patients who had survived through treatment for CCM, it continued to improve over the 6 months following diagnosis. Low weight at diagnosis, prolonged hospital admission, and being Asian were associated with lower QOL. QOL is an important outcome that should be considered among HIV infected patients treated for serious infections such as CCM.

## Introduction

Cryptococcal meningitis (CCM) is one of the leading causes of mortality among HIV infected patients, especially in sub-Saharan Africa and Asia. Mortality from CCM remains high, in spite of a significant reduction in the incidence of opportunistic infections (OIs), resulting from improvement in HIV management including early initiation of antiretroviral therapy (ART) [1]. Between 2009 and 2014, the estimated global annual prevalence and mortality due to CCM reduced from 975,900 to 223,100 and 624,700 to 181,100 respectively [2,3]. While incidence, prevalence, and mortality indices have been used widely to demonstrate the burden of CCM amongst HIV infected patients, its effects on other important measures of health such as quality of life (QOL) have not been adequately evaluated. Due to the advanced stage of AIDS at which CCM occurs, severity of disease pathology, toxicity of antifungal medications and disease sequelae, the QOL post-treatment is likely to be poor. We previously showed that patients with low QOL at treatment completion are more likely to die two years later than those with higher QOL[4]. Previous research has demonstrated that QOL measurements amongst HIV infected patients should be done routinely [5-8], since improved QOL is one of the ultimate goals of HIV management. We used data collected from patients enrolled in the CryptoDex trial (PMID: 25391338, ISRCTN59144167) [9,10] to determine QOL at 10 weeks and six months since treatment initiation and the factors associated with QOL at 10 weeks and six months.

## Methodology

### Study design and setting

The CryptoDex trial was a double-blind placebo-controlled phase III trial of adjunctive dexamethasone in HIV infected adults with cryptococcal meningitis. The study was conducted between 2013 and 2015 at thirteen sites in six countries in Asia and Africa (Vietnam, Thailand, Indonesia, Laos, Uganda, and Malawi).

### Subjects enrolled in the CryptoDex study and their management

Patients recruited in the CryptoDex study were 18 years and above, had HIV infection, a clinical syndrome consistent with CCM, and microbiological confirmation of disease, as indicated by one or more of the following test results: 1) positive India ink staining of cerebral spinal fluid (CSF); 2) culture of *Cryptococcus* species from CSF or blood; or 3) cryptococcal antigen detected in CSF on cryptococcal antigen lateral flow assay (IMMY). Exclusion criteria included pregnancy, renal failure, gastrointestinal bleeding, having been treated with more than 7 days of anticryptococcal antifungal therapy, and being on or requiring glucocorticoid therapy for coexisting conditions. Informed consent was obtained from all patients or a next of kin where a patient was not in position to consent. Eligible patients were randomised in a 1:1 ratio, stratified by site, to receive either dexamethasone adjunctive therapy or placebo in a tapered dose until 42 days post-randomisation. Details of other study procedures and investigations can be found in the CryptoDex study protocol [11]. The treatment regimen included an induction phase with intravenous Amphotericin B (1 mg/kg/day) and oral Fluconazole 800mg daily for 14 days, followed by a continuation phase of 800mg daily fluconazole for eight weeks, and then a maintenance phase with oral fluconazole 200mg daily. This regimen was consistent with the WHO treatment recommendation for settings where Flucytosine was not available at the time the trial was conducted [12].

Patients were followed up in the study until six months after enrolment, at which point they were exited and referred back to their HIV care clinics for continuation of HIV care.

### Subjects included in this analysis

This QOL analysis includes patients who survived up to 10 weeks post-treatment initiation when the first QOL assessment was performed. A second QOL assessment was done at month six and therefore patients who had not died between 10 weeks and six months and were accessible i.e., able to come to the clinics, are included in the six months analysis.

### Variables and their measurement in the CryptoDex study

Quality of life was measured using the EQ-5D-3L tool [13]. This tool gives a subjective measure for the QOL, which is in line with the definition of QOL by WHO. The WHO defines QOL as an individuals’ perception of their position in the context of culture and value systems in which they live and in relation to their goals, expectations, standards, and concerns [14]. The tool is a widely used measure of health status consisting of two parts. The first part (the descriptive system) assesses health in five dimensions (mobility, self-care, usual activities, pain/discomfort, and anxiety/depression). Each dimension has three levels of response (no problems, some problems, extreme problems/not able to). This part of the EQ-5D-3L questionnaire provides a descriptive profile that can be used to generate a health state profile. For example, a patient in health state 11233 would have no problems in mobility and self-care, some problems with usual activities, severe/extreme pain/discomfort, and extreme anxiety or depression. Each health state is assigned a summary index score based on societal preference weights for the health state. Heath state index scores generally range from less than 0 (where 0 is a health state equivalent to death; negative values are valued as worse than death) to 1 (perfect health). Health state preferences can differ between countries. Where a value set is not available for a country/region, the assessor can opt to select a value set for a country/region considered to most closely approximate the country of interest [13]. We used the Thailand value set for sites in Asia (Vietnam, Thailand, Indonesia and Laos) [15] and the Zimbabwe value set for sites in Africa (Uganda and Malawi) [16]. The second part of the EQ-5D-3L questionnaire consists of a visual analog scale (VAS) on which the patient rates his/her self-perceived health from 0 (the worst imaginable health) to 100 (the best imaginable health/perfect health).

The tool was translated into the local languages and administered by a study doctor who read the questions to the patient. Other details concerning its administration can be found in the study protocol [11].

We measured the impact of a number of variables on QOL measured using the VAS score including: demographic characteristics (age, sex, and education measured as number of years in school); continent, i.e. Asia or Africa; ART status, i.e. on or not on ART; time on ART; most recent CD4 count at enrolment; presence of convulsions; intervention arm, i.e., dexamethasone or placebo; Number of inpatient days by week 10 and month six, baseline CSF fungal burden, and Number of adverse events (neurological events, new AIDs defining illnesses, other adverse events, and immune reconstitution syndrome events).

### Data Analysis

All data from the CryptoDex study was derived from the secure OUCRU proprietary CLiRES trial database. Data were extracted into a Microsoft Access database for export to STATA (College Station, TX, version 15.0). Categorical variables were summarised by frequencies and percentages, while continuous variables were summarised using means (standard deviations) and/or medians (interquartile ranges).

A health profile was generated by continent and time points (i.e., week 10 and month six), from which we derived a health state index score using the selected value sets for each region. We then summarised the index scores by means, standard deviations (SD), and minimum and maximum scores by continent at the different time points. VAS scores were interpreted as very good (81-100), good (51-80), normal (31-50) and bad/very bad (0-30) [17]. T-tests were used to compare VAS scores between continents and to test for significant changes in VAS scores and index scores at the different time points. We also computed and presented percentages of categories under each of the five dimensions of the EQ-5D-3L questionnaire.

We used simple linear regression to determine crude associations between VAS scores at week 10 and month six and various variables. This analysis was not repeated using index scores as two value sets (Thailand and Zimbabwe) were used, and sample sizes were too small to conduct separate analyses. Variables that had a p-value of 0.2 and below at univariate analysis were included in a multivariable linear regression model. Variables were retained in the multivariable model if their inclusion did not make the model significantly worse at LRT p-value 0.05.

### Ethical considerations

The CryptoDex study received ethical approval from each country’s respective regulatory authorities, and the Oxford Tropical Ethics Committee as indicated in the published protocol and study paper [9,10].

## Results

### Baseline Characteristics

Figure 1 is a flow diagram showing the patients included in this analysis. Out of 823 patients screened at the 13 sites globally, 451 were found eligible and randomized. One patient was subsequently excluded because he never received the assigned study drug due to an administrative error. Out of the remaining 450 patients, 251 (55.8%) survived to week 10 post-treatment initiation, and 238 (94.8%) completed a QOL assessment and were included in our analysis. The other 13 (5.2%) patients were not accessible for the quality of life assessment since they did not come to the clinics. At baseline, the mean age (SD) of the 238 patients was 35.2 (8.5) years, and 101 (42.4%) were on ART. The median CD4 count (IQR) was 30 (13, 71) cells/ml. Thirty (12.6%) patients reported a history of convulsions. Of the 238 patients, 113 (47.5%) had been randomised to receive dexamethasone adjunctive therapy. The baseline characteristics are summarised in Table 1.

**Table 1:**
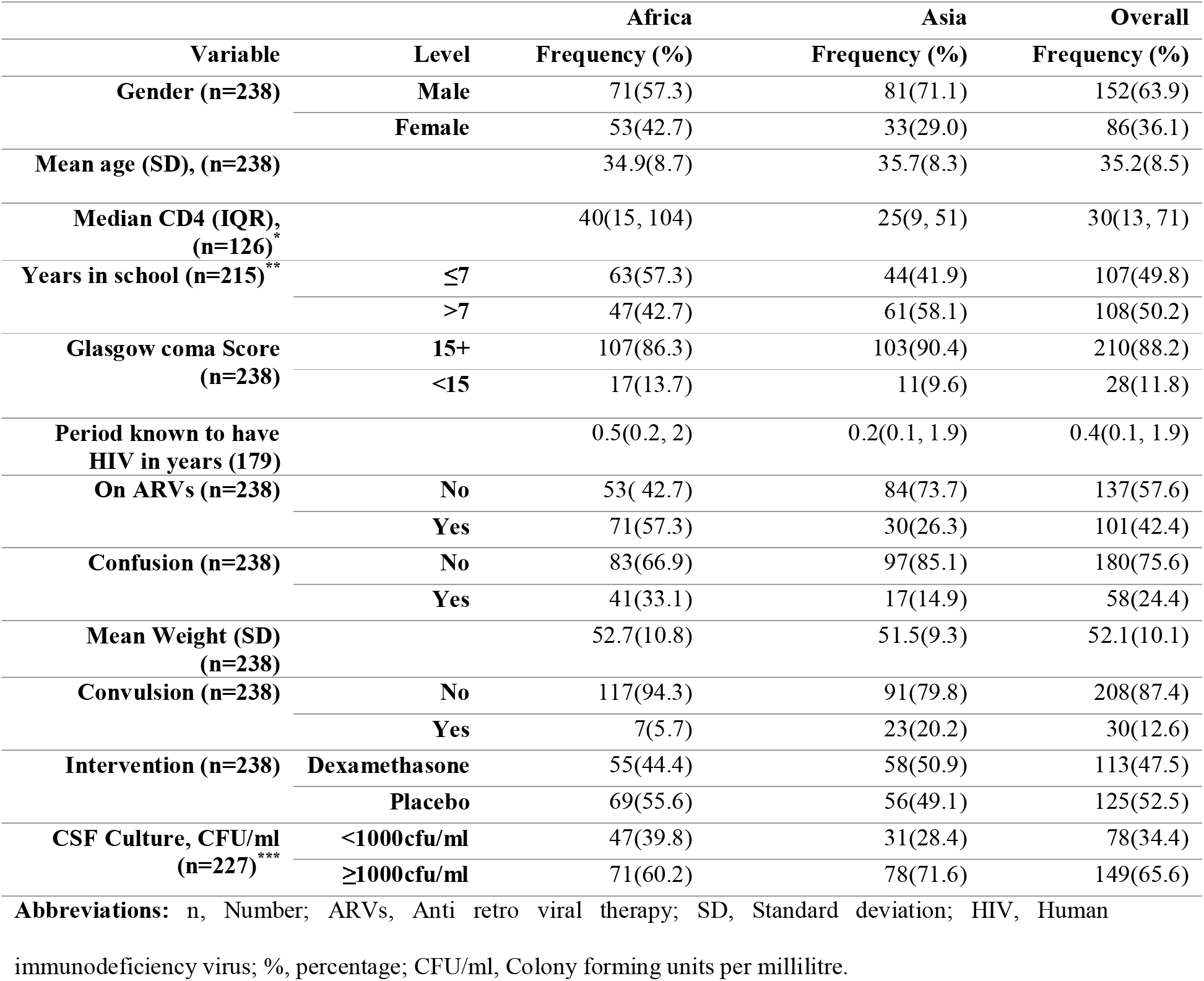
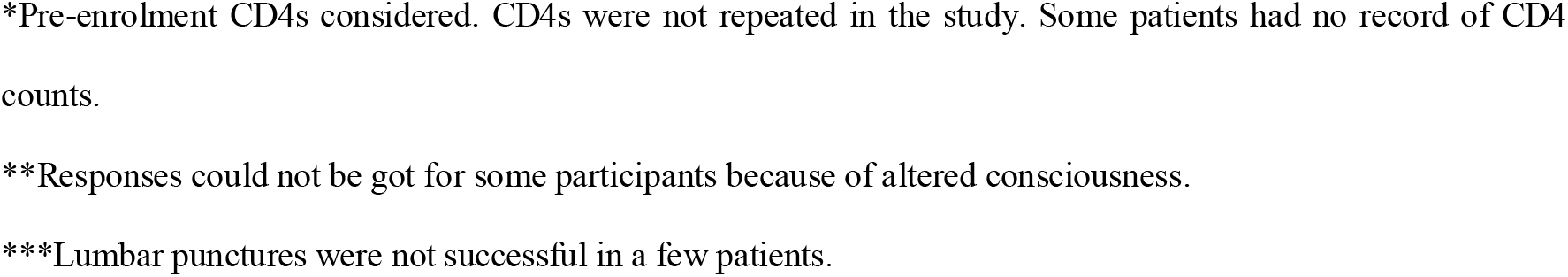
Baseline characteristics of the patients that had QOL evaluation at 10 weeks.

**Fig 1:**
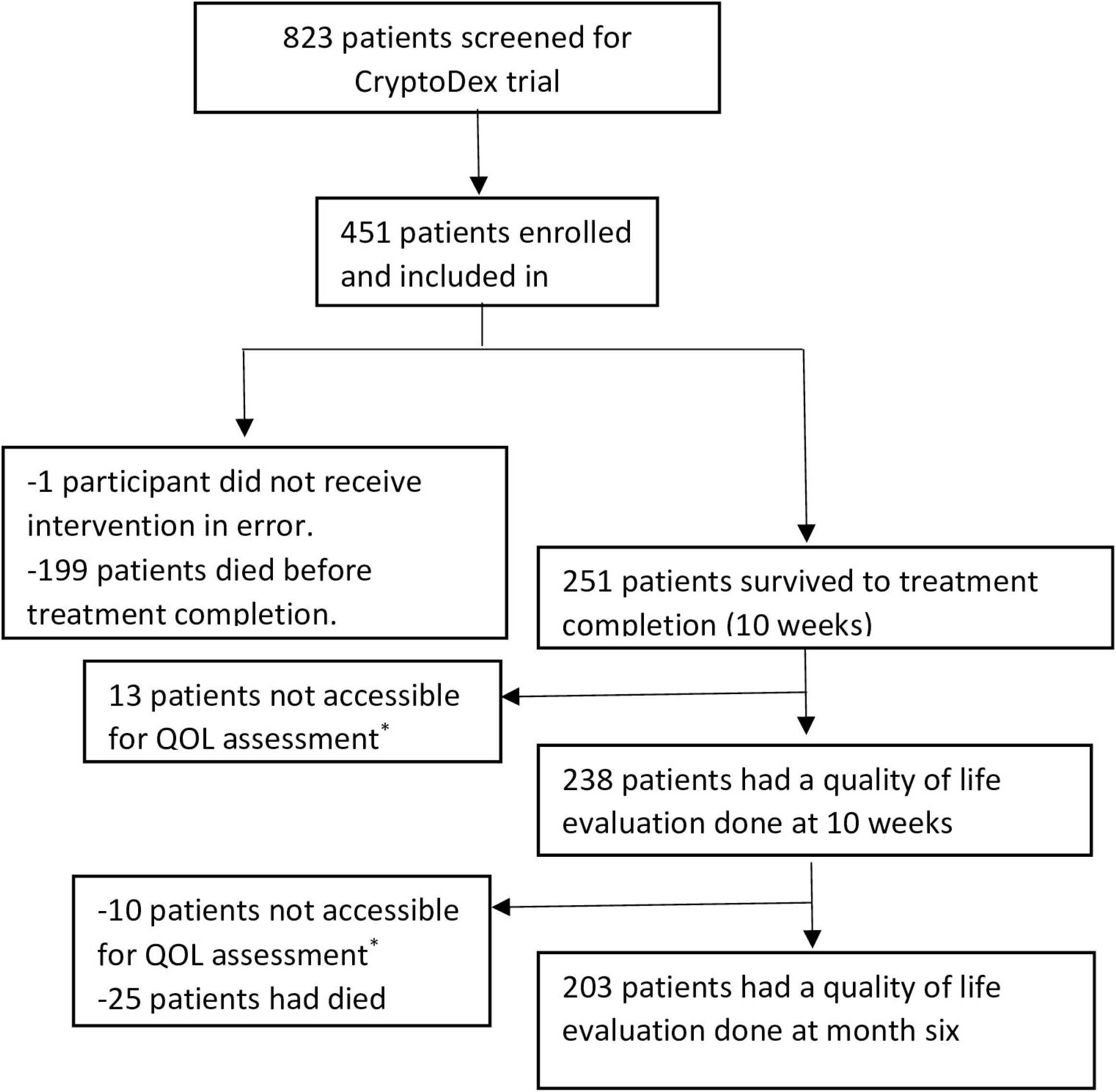
Flow diagram showing participants enrolled in CryptoDex trial who had quality of life evaluations done. * Patients were not able to come to clinics to have QOL assessment.

### Adverse events

Four hundred and thirty four adverse events were captured among these participants, including 67 new neurological events, 93 new AIDs defining illnesses, 365 other adverse events, and 9 immune reconstitution syndrome events.

### Quality of life assessment

#### Descriptive system and index scores

Health profile results for patients at both time points are summarised in Table 2. There was no difference in the proportion of patients from Africa or Asia rating themselves as having perfect health (i.e., profile 11111) at 10 weeks (Africa: 40 of 124 (32.3%); Asia: 37 of 114 (32.5%)). At this time point, 4 Asian patients (3.5%) rated themselves as having the worst health state possible i.e. 33333 compared with 1 African patient (0.8%, p=0.2). At month six, reported quality of life had increased in both African and Asian centres, with 58 of 104 (55.8%) Africans and 48 of 99 (48.5%) Asians rating themselves as having perfect health (p=0.2).

**Table 2:**
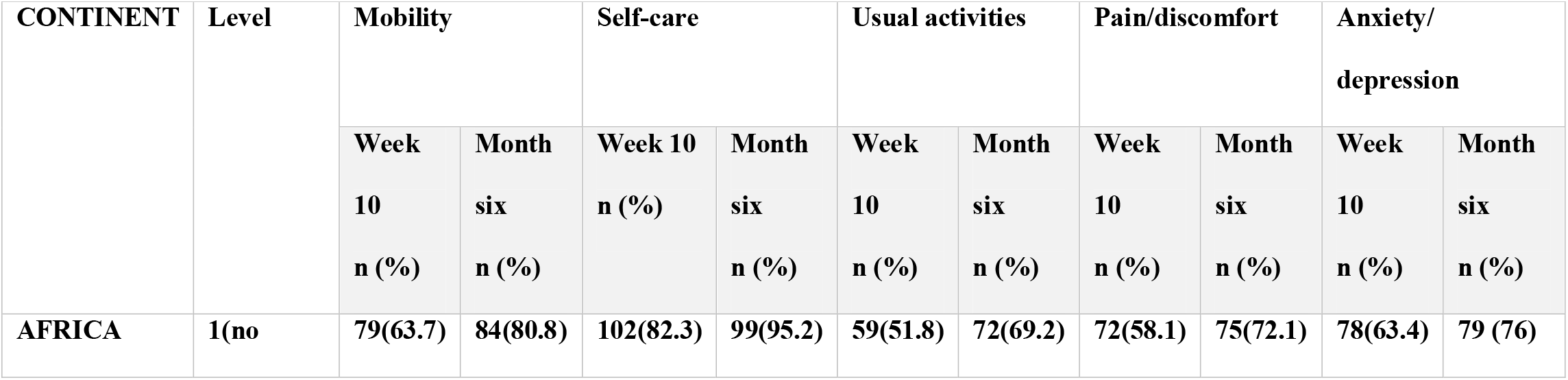
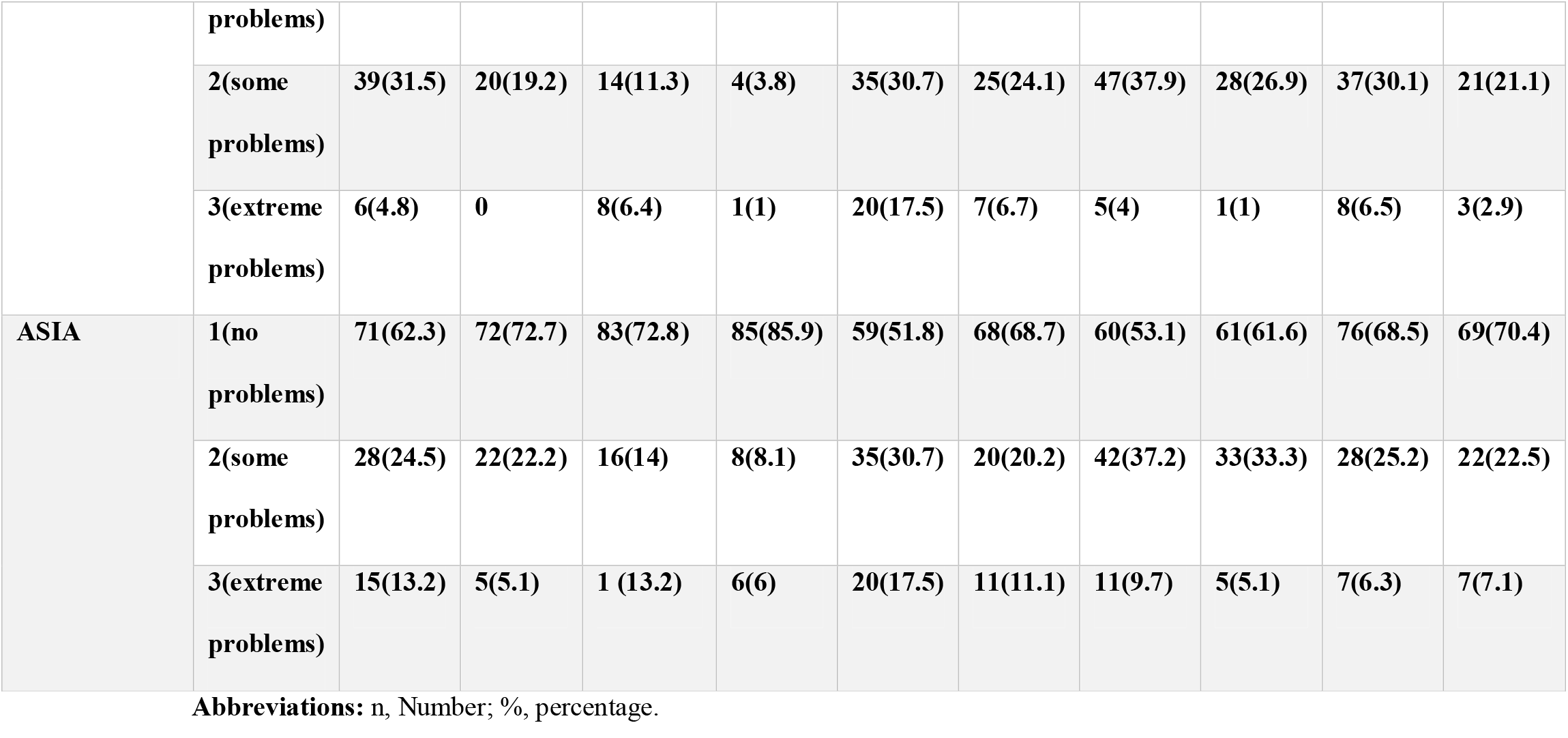
Health profile scores for patients at week 10 and month six.

At week 10, the mean index scores (SD, range) were 0.785 (0.2, -0.145 to 1) and 0.619 (0.4, - 0.452 to 1) for African and Asian patients respectively, increasing significantly at month six, to 0.879 (0.2, 0.341 to 1), p=0.002 and 0.731 (0.4, -0.452 to 1), p=0.052 for African and Asian patients respectively.

### EQ VAS scale results

The overall mean VAS score (SD) at 10 weeks was 57.2 (29.7), increasing significantly to 72 (27.4) at month six (p<0.001), both mean scores falling within category 51-80, i.e. “Good QOL”. The mean score (SD) at 10 weeks was significantly higher among African patients than Asian patients i.e., 65.7 (21.5) vs 47.5 (34.5), p<0.001. Similarly the mean score (SD) at six months was significantly higher among African patients compared to Asian patients i.e., 79 (17.7) vs 64.2 (33.4), p<0.001.

Mean VAS and Index scores at week 10 and month six are represented in Figure 2.

**Fig 2:**
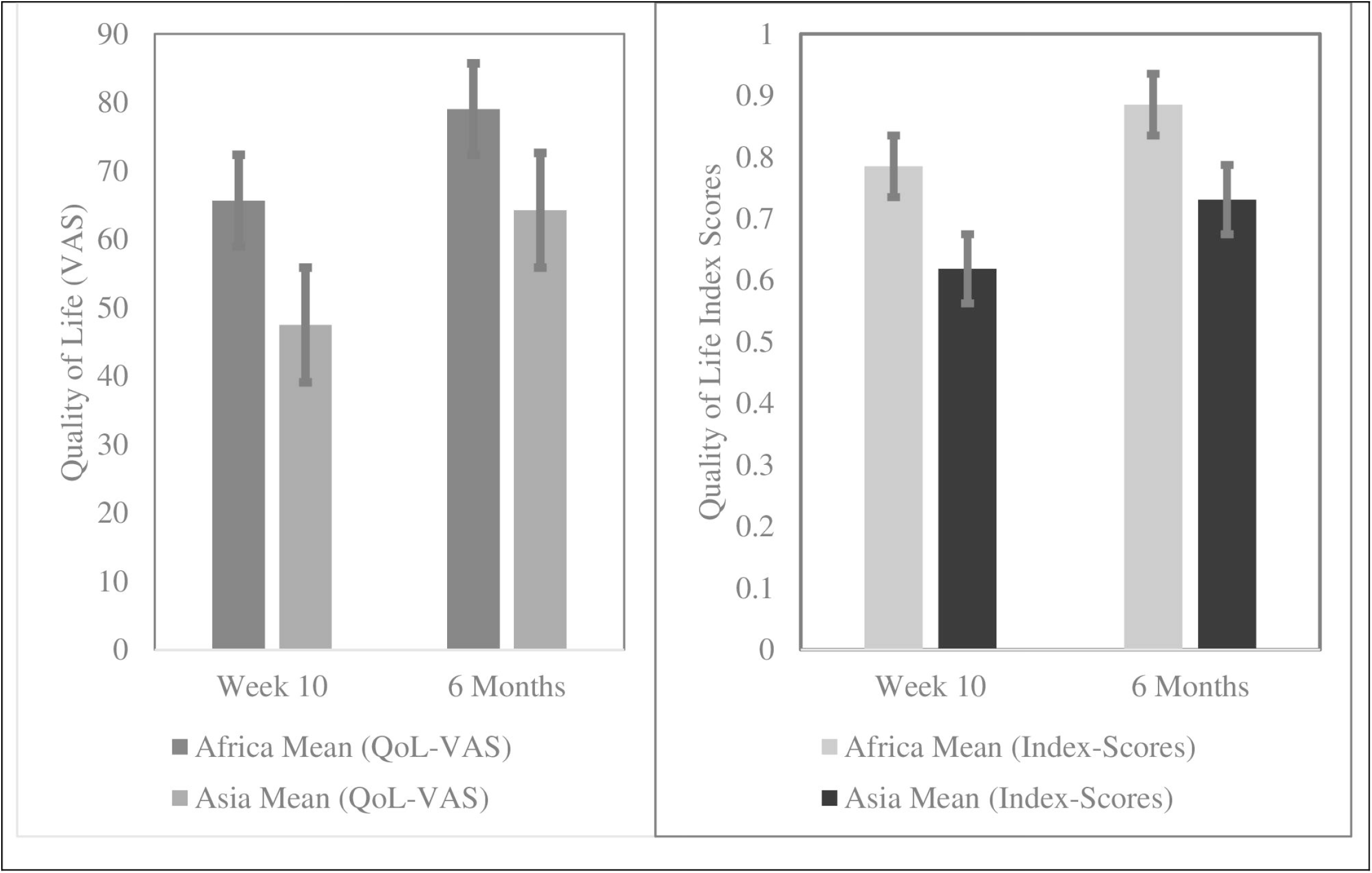
Mean VAS scores and Index scores at week 10 and 6 months after treatment initiation. Abbreviations: VAS, Visual analog scale; QoL, Quality of life.

### Factors associated with quality of life

Results from the univariate and multivariate analysis are summarised in Table 3.

**Table 3:**
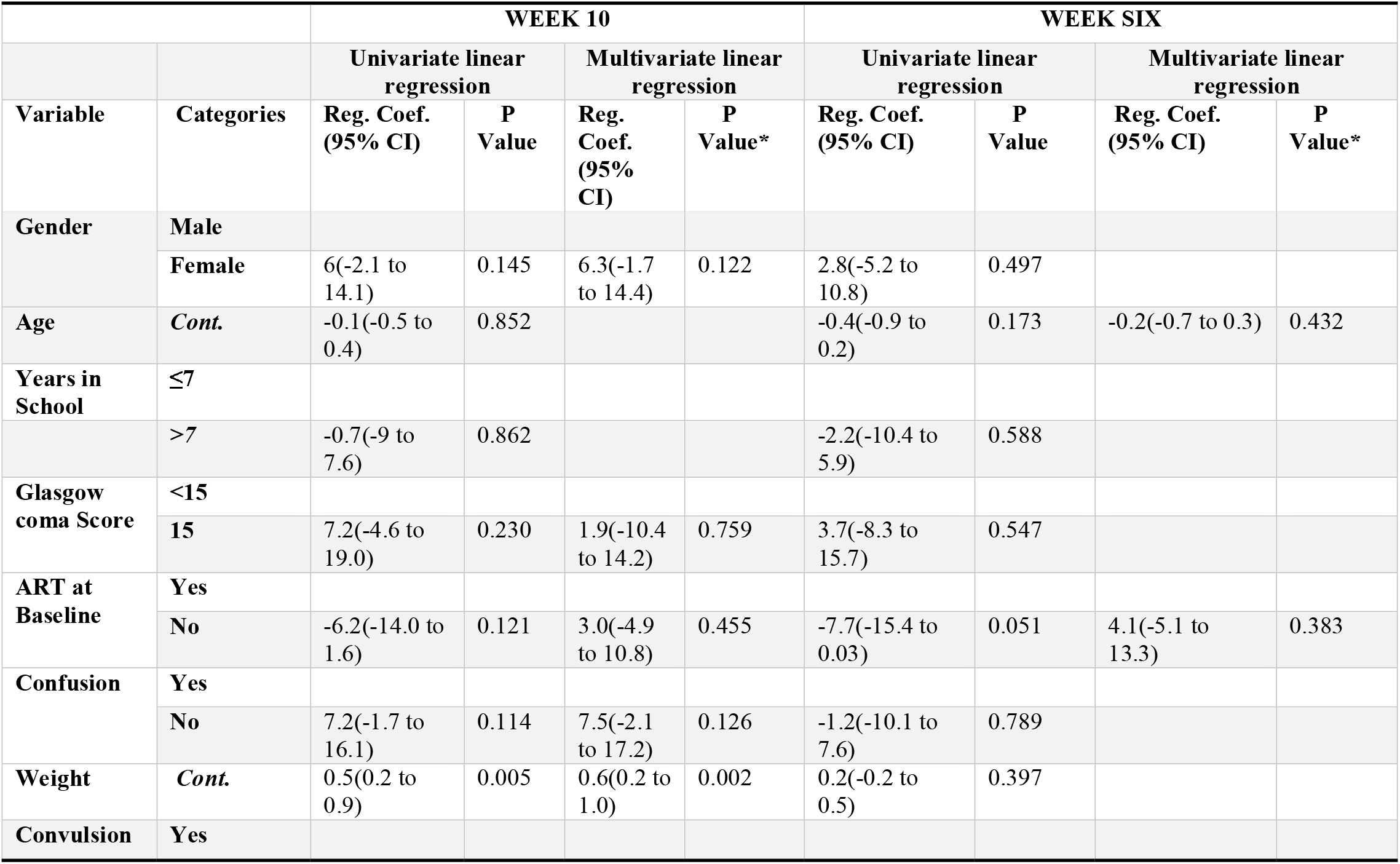
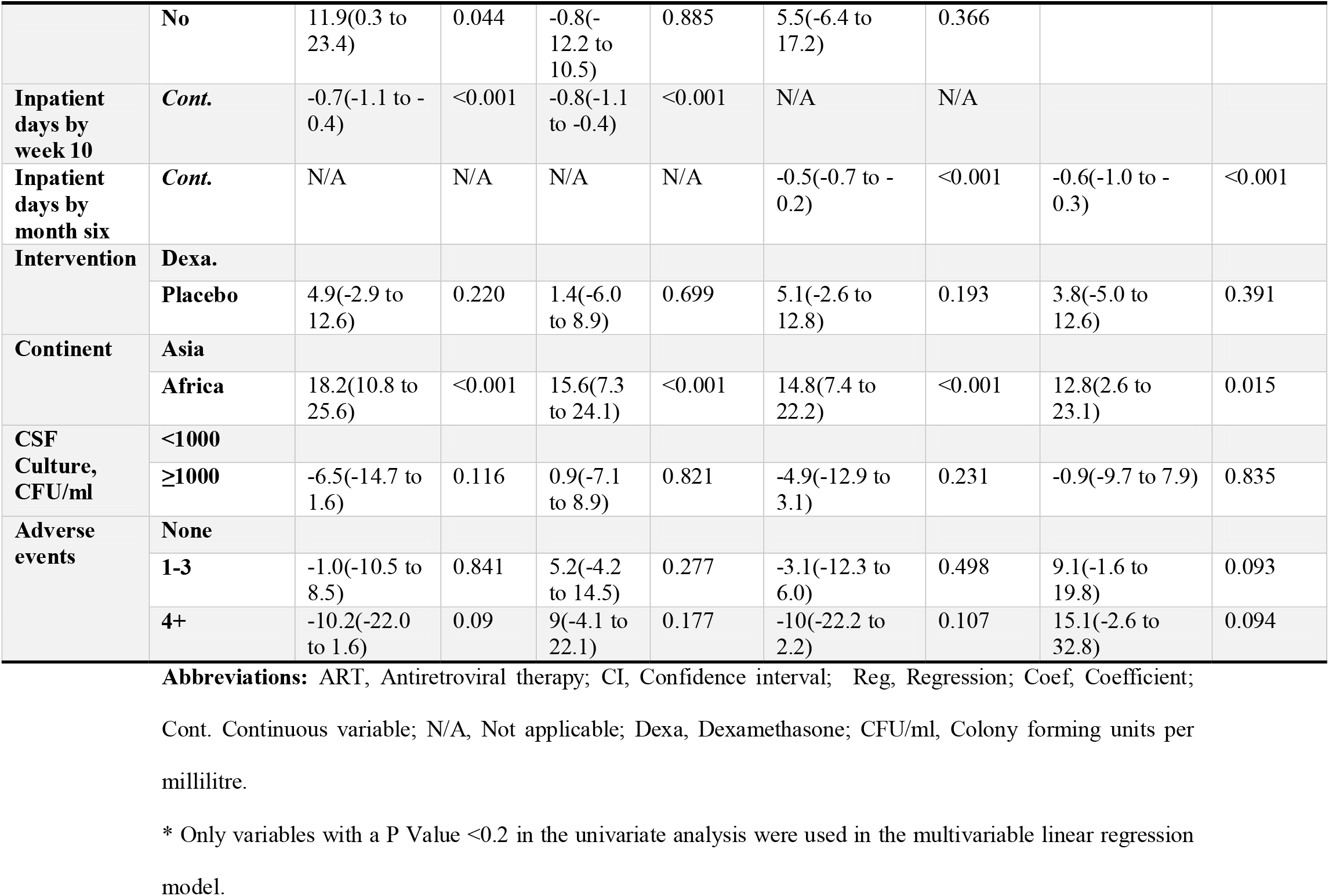
Quality of Life and associated factors at 10 weeks and 6 months.

At week 10, the baseline factors associated with higher VAS score at multivariate analysis were: greater weight (regression coefficient 0.6, 95% CI: 0.2 to 1), p=0.002; and being from Africa (regression coefficient 15.6, 95% CI: 7.3 to 24.1), p<0.001. Higher inpatient days by week 10 was associated with poorer VAS score (regression coefficient -0.8, 95% CI -1.1 to - 0.4), p<0.001.

At month six, the only factor associated with higher VAS score at multivariate analysis was African origin (regression coefficient 12.8 95% CI 2.6 to 23.1), p=0.015. Again, higher inpatient days was associated with a reduced VAS score at 6 months (regression coefficient - 0.6, 95% CI -1.0 to -0.3, p=<0.001).

## Discussion

HIV-associated CCM is an extremely debilitating disease associated with high mortality. Affected patients present with advanced HIV disease and considerable immunosuppression. While the disease itself is associated with significant risks of sequelae including neurological deficits, the treatment itself, of which amphotericin is the backbone, is also associated with significant toxicities [18,19]. All these factors have the potential to reduce the QOL among patients who survive up to treatment completion.

We were pleased to see that many patients reported relatively good self-perceived QOL following completion of treatment for CCM, and that this tended to improve thereafter through the 6 months following diagnosis. However, given the nature of the disease and its treatment, it is perhaps not surprising that very few patients reported ‘very good’ or ‘perfect’ health. We did not assess the QOL at study entry in the CryptoDex trial and thus could not measure improvement directly attributable to antifungal treatment. However, we found that patients reported significant improvements in QOL between 10 weeks and six months after diagnosis, both with the descriptive and VAS scales. These continuing improvements in QOL are likely due to multiple factors. These may include gradual improvement due to neurological recovery, better engagement with health services, and the use of antiretroviral drugs. A previous study identified that patients with CCM had significant improvements in QOL compared with pre-treatment scores [5]. It has been shown previously that different antifungal combinations can be associated with long term (6 month) differences in survival[9]. Therefore, while some of the improvement in QOL seen in our patients, who all received the same antifungal treatment, may be due to the factors we described above, developing more effective induction and consolidation therapy for CCM is likely to yield further improvements in QOL.

We consider QOL an important health parameter to measure since better scores have been shown to correlate with long term survival [4,20]. In our study, we used the EQ-5D-3L tool. This has the advantages of simplicity and ease of administration including amongst very sick patients, as well as ease of scoring and interpretation. While it has not been used previously in many studies of CCM patients, our work shows that it can identify changes in QOL over time following treatment of CCM. As such, it has the potential to identify patients who may benefit from increased follow-up during the recovery process – further studies are needed. Identifying such patients could help with distribution of scarce resources and ensure the best possible outcomes for all patients.

We found that prolonged admission by week 10 and month 6 were associated with lower QOL measured using the VAS at week 10 and month six. This is not surprising – patients admitted for longer periods were likely more ill at baseline. A similar finding was reported in a study done by Nafteux and colleagues in Belgium [21].

We were surprised to see that patients from African countries in the CryptoDex trial rated themselves to have better QOL at week 10 and six months than did Asian patients. This effect was seen with both the descriptive scale and VAS. This effect remained apparent even after adjustment for other factors that affected QOL. We suggest that this is probably a societal perceptual variation rather than an indication of impact of disease, although its inverse correlation with the gross national income per capita for the Asian and African countries was counter-intuitive. We also found that higher weight was associated with higher VAS at week 10. The most likely explanation here is that low weight is correlated with more severe disease [22]. Unlike a previous study from Uganda amongst HIV patients that found male gender was associated with better quality of life, we did not identify a difference between males and females in terms of QOL following treatment for CCM [23].

Our analysis has some limitations. First, we used a subjective measure of QOL, which may not be an absolute indication of the QOL of an individual. Naturally, optimistic people may rate their health status higher than pessimistic ones. However, the WHO recommends that QOL tools should measure from the subjective viewpoint since the patients experience is more relevant than the assessment of an objective observer [24]. Second, we used health index scores derived from models of countries that we assumed were similar to our own, since models do not exist for all the countries in our study. Use of validated index scores from the actual countries that participated in CryptoDex could conceivably result in different outcomes. However, we think this is unlikely because we measured QOL at two time points and found improvement over time.

Going forward, we would recommend that QOL assessment should be part of all trials in cryptococcal meningitis. Recent work from Molloy and colleagues has suggested that current doses of amphotericin, which clearly have the greatest antifungal effect, may in fact be resulting in patient harm because of associated toxicities [25]. QOL measurements among survivors could help in parsing such data, while we wait for more effective and tolerable novel treatments to be developed.

## Conclusion

While self-perceived QOL was only relatively good among this cohort of patients who had survived through treatment for CCM, it continued to improve over the 6 months following diagnosis. Low weight at diagnosis, prolonged hospital admission, and being Asian were associated with lower perceived QOL. QOL is an important outcome that should be considered among HIV infected patients treated for serious infections such as CCM.

## Data Availability

Data used for this paper will not be shared publically, but if there is need, the data can be retrieved following the data sharing policy at the MRC/UVRI & LSHTM Uganda Research Unit. Tis policy can be assessed through the following link: https://www.mrcuganda.org/publications/data-sharing-policy

## Acknowledgements

We would like to appreciate the team that implemented the CryptoDex trial at all the sites in Africa and Asia.

We are especially grateful to the study participants and/or their representatives for the precious time they accorded us.

The content is solely the responsibility of the authors and does not necessarily represent the official views of the supporting agencies.

## Author Contributions

Conceived and designed the experiments: Jonathan Kitonsa. Performed the experiments: Jonathan Kitonsa, Freddie Kibengo and Zacchaeus Anywaine. Analysed the data: Julius Kiwanuka, Sheila Katusiime, and Jonathan Kitonsa. Wrote the paper: Jonathan Kitonsa, Julius Kiwanuka, Freddie Kibengo, Kenneth Katumba, Zacchaeus Anywaine, Pontiano Kaleebu, Alastair Gray, and Jeremy Day.

